# Soundscapes in acute and rehabilitation NHS hospital environments: a mixed methods study

**DOI:** 10.1101/2025.07.01.25330619

**Authors:** Fiona Marshall, Peter Rutherford, Hannah Elliott, Olivia Smith, David Baguley, Laura Edwards

## Abstract

**Objectives:** To assess hospital soundscapes, typically within those departments where patients with Acquired Brain Injury (ABI) are cared for. To investigate soundscapes’ effects among staff and patients.

**Design:** A mixed methods observational study

**Setting:** 5 units within an NHS hospital Trust

**Participants:** Patients with and without ABI (n=31); staff members (n=23)

**Interventions:** n/a

**Outcome measures:** Sound level measurements were collected over 7-day periods; interviews and questionnaires were conducted with staff and patients. Qualitative and quantitative data were triangulated.

**Results:** Sound pressure levels consistently exceeded World Health Organization recommendations. Higher levels and unpredictable/irregular sounds were perceived as most disruptive to staff and patients. Escalations of sound affected ease and effectiveness of communication, further exacerbated by use of personal protective equipment. Patients tended to be more tolerant of disruptive sounds during the day but less so at night. Intermittent equipment sounds, sounds of patient agitation, and instances of staff discussing inappropriate topics for a hospital setting negatively affected both patients and staff. Certain sounds were perceived positively and helped patients, particularly those with ABI, orient themselves in their surroundings.

**Conclusions:** Soundscapes in hospital settings are important for patient and staff health and wellbeing. Recommendations towards improved sound environments are provided.

## Introduction

Hospital noises and soundscapes have been recognised for over 100 years to be important to patients and staff(1, 2). “Soundscape” is a term used to describe the sounds in an environment and the effects they have on a listener(3). It recognises the importance of individual experience in responding to sensory input - for instance, one patient may find that a radio playing music is distracting and pleasant during their hospital stay, while another may find the identical sound intrusive and annoying.

Sound and noise in hospital have been previously shown to affect sleep and mood in hospital inpatients(4). Individuals with acquired brain injury (ABI) may experience sounds as more intrusive, even when sound measurements indicate normal sound pressure levels(5). Sleep disturbance is very common following ABI, compounded by increased sleep requirements, disrupted circadian cycles and heightened noise sensitivity(6, 7). Patients with ABI often receive care across multiple units, in places such as intensive care units and emergency departments, which have been shown to be loud and disorientating environments, yet the impact of sound on the care experience of people with ABI is largely undescribed in the literature.

Many hospital staff work long shifts more than 8 hours(8), and longer periods exposed to high sound levels may lead to chronic health and well-being issues, hearing loss and inherent stress(9). Staff contend with multiple concurrent sound sources, and over time, the myriads of sounds can impact on staff wellbeing, contributing towards job dissatisfaction, burnout and performance issues(10).

The COVID-19 pandemic resulted in widespread introduction of personal protective equipment, reduced hospital visiting and social distancing precautions. These factors all represent significant potential changes to communication, and subsequently to the soundscape(11).

To understand how soundscapes might influence patient experiences for staff and patients with and without ABI, we conducted a mixed-method study across several hospital units, including sound measurements, questionnaires and interviews.

Uniquely, we were able to collect data during the COVID-19 pandemic.

### Aims

To explore the possible impacts of soundscapes on patient and staff experience in five wards and departments where people with acquired brain injury commonly receive care: Emergency Department; Medical Assessment Unit; Adult Intensive Care Unit; Hyperacute Stroke Unit; Neurorehabilitation Unit.

### Objectives

- To measure sound pressure levels in these wards and departments and compare to established international guidelines,
- To assess staff and patient expectations, experiences, and perceptions of soundscapes on these units using questionnaires and interviews,
- To integrate these findings, understand their impact and provide recommendations on soundscape management and mitigation.

## Methods

### Study design and setting

We conducted a hospital-based mixed methods study, combining qualitative and quantitative data, informed by patient and public consultation.

Figure 1 outlines the study design.

**Figure 1:**
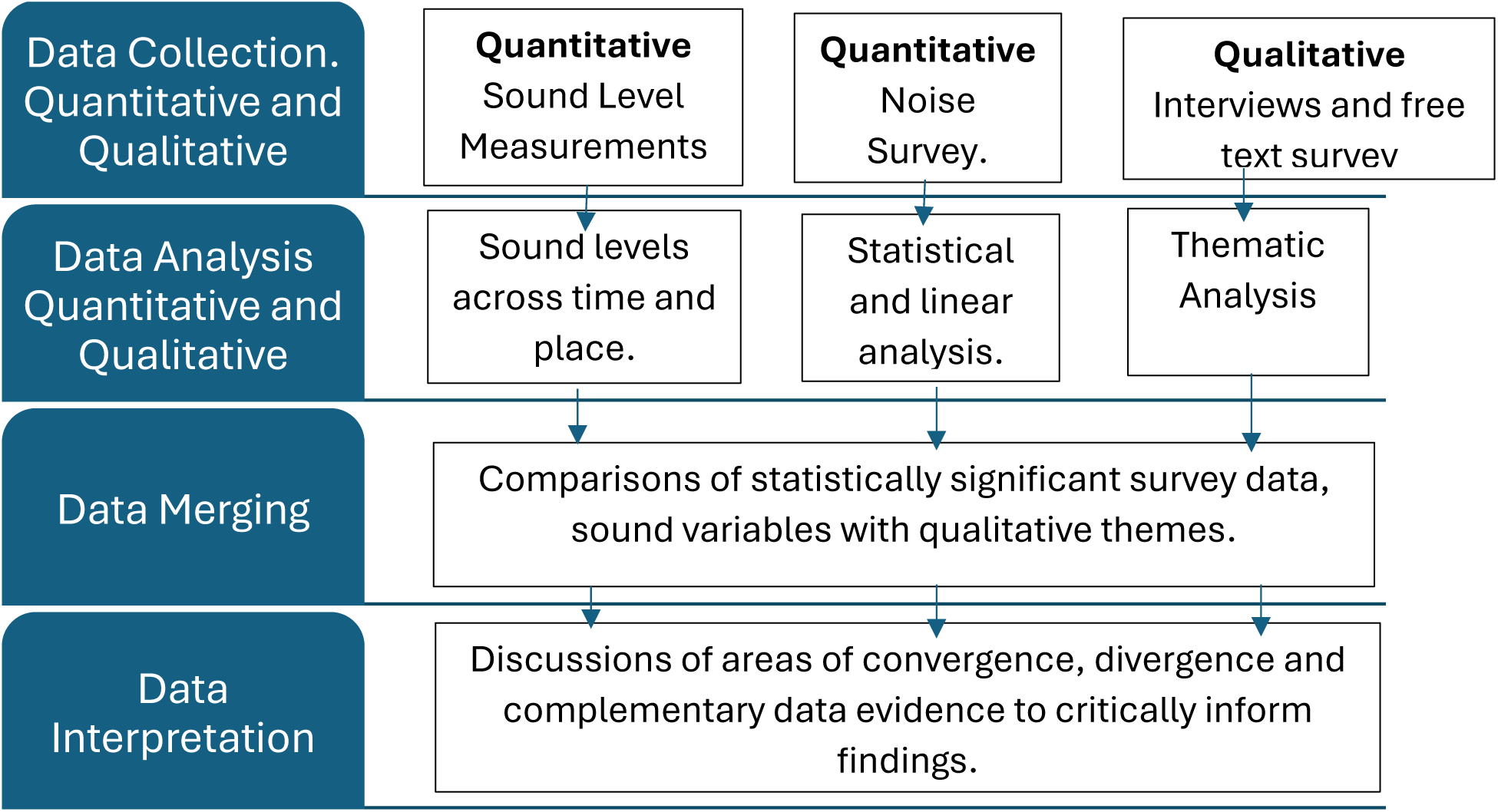
Flow chart demonstrating data collection, analysis, interpretation and triangulation

The study was conducted at the two Derby sites: the Royal Derby Hospital (RDH) and the Florence Nightingale Community Hospital (FNCH), both part of the University Hospitals of Derby and Burton NHS Foundation Trust. Specifically, sound level measurements and participant recruitment took place in Emergency Department (ED), Medical Assessment Unit (MAU), Adult Intensive Care Unit (AICU), Hyperacute Stroke Unit (HASU), all at RDH, and Neurorehabilitation Unit (NRU) at FNCH. These locations were selected based on the typical patient pathway for individuals admitted with acquired brain injury.

The study was conducted between November 2020 and August 2022. All sound level measurements were conducted between November 2020 and November 2021, with 50% of participants recruited before November 2021, i.e. during periods of restricted hospital visiting due to the COVID-19 pandemic.

## Sound Data Collection

### Sound levels

Sound levels were recorded for 7 consecutive days and nights in each of the 5 units using a Trojan 2 Noise Nuisance Recorder from Cirrus Research plc, a Class 1 Sound Level Meter. Recorder locations were chosen after discussions with staff members to ensure they provided an accurate representation of the sound levels experienced by ward inhabitants, without compromising the safety of ward operations. The location in MAU was furthest from day-to-day patient activity, being set in an alcove 2.8 metres from a busy main corridor and 6.1 metres from the nearest patient bed. The remainder of the locations were less than 2 metres from the nearest patient bed. The positioning of the sound level meter in relation to the patient bed was not standardised across all settings as it was unreasonable to secure equipment in such a fast-paced environment, where demand for staff resources including maintenance staff was higher than usual.

The sound level meter was calibrated before and after each measurement period using a Cirrus Research Class 1 calibrator. Sound pressure levels were logged every second resulting in 604,800 discrete measurements over each 7-day period. For analysis purposes, these were subsequently reduced to 168 x 1-hour LA_eq_ measurements.

### Quantitative Sound Measurement Analysis

Data were analysed using Cirrus Research NoiseTools software (Cirrus Research UK) and Microsoft Excel. Acoustic metrics were compared to WHO guidelines(12) for acceptable sound levels in hospital settings (Table 1). In addition to LA_eq_ and LA_max_, the analyses included derived measurements of sleep quality and annoyance.

**Table 1:** Guidelines and measurements used for reference through this study; variables derived from sound level recordings. WHO = World Health Organisation; dB(A) = A-weighted decibels.

| Measurement | Meaning | WHO guidelines(12) |
| --- | --- | --- |
| $LA_{eq}$ Day | Equivalent sound pressure level ($LA_{eq}$) measured from 07:00-19:00 | No greater than 35dB(A) in rooms where patients are being treated or observed (i.e. not sleeping)<br>No greater than 30dB(A) in rooms where patients are sleeping |
| $LA_{eq}$ Evening | $LA_{eq}$ measured between 19:00-23:00 | N/A |
| $LA_{eq}$ Night | $LA_{eq}$ measured between 23:00-07:00 | No greater than 30dB(A) |
| $LA_{max}$ Night | Maximum sound pressure level | No greater than 40dB(A). Maximum single event of 45dB(A) |
|  | measured between<br>23:00-07:00 |  |
| $L_{den}$ | Average sound pressure over day, evening and night | Descriptor of noise level with 5dB(A) penalty imposed for evening and 10dB(A) for night time noise |
| **Metric** | **Assessment protocol** |  |
| Annoyance | The propensity to cause annoyance (defined here as the sensation of displeasure arising from key attributes of the acoustic environment), and its severity can be mapped to background sound pressure levels according to WHO guidelines (12) as follows:<br>• > 55dB $LA_{eq}$, “Serious annoyance”<br>• > 50, < 55db $LA_{eq}$, “Moderate annoyance”<br>“Sound pressure levels during the evening and night should be 5 – 10 dB lower than during the day.....it is emphasized that for intermittent noise, it is necessary to take into account the maximum sound pressure level as well as the number of noise events.”(12) |  |
| Sleep | Sound levels with the propensity to disturb sleep are defined by the WHO community noise guidelines(12) as:<br>• < 30dB $LA_{eq}$ likely to result in good sleep quality.<br>• < 35dB $LA_{eq}$ likely to result in reasonable sleep quality.<br>< 40dB $LA_{max}$ and should not be exceeded 10 to 15x per night to reduce interruption. |  |
| Sound sensitivity by time of day | $L_{den}$ is the equivalent sound pressure level day, evening night). This imposes a sound pressure level penalty of 5dB(A) and 10dB(A) for the evening and night periods respectively, representing our increased sensitivity to sound during these periods. These were compared to the daily $LA_{eq}$ (which is the equivalent sound pressure level measured over the 24 hour period). Statistical analyses were performed to the hourly $LA_{eq}$ data. These analyses were used to gauge the percentage of time that a particular sound pressure level was exceeded in the ward. |  |

### Participant selection

Staff members and patients were recruited by a clinical staff member (LE) from each of the selected units. This included inpatients with and without ABI (to gain perspectives of people who could communicate freely and who were sharing their care settings with people with ABI), as well as those who had been treated for ABI and subsequently discharged.

Inclusion criteria for both patients and staff required participants to be aged 18 years or over and being capable of providing informed consent. Staff members were eligible if they had been working as permanent staff on the unit for at least 4 weeks. Patients were eligible at any point during their stay in ED or MAU and after a 48-hour stay in other units.

### Qualitative data collection

Semi-structured interviews were conducted with participants by a consultant clinician (LE) and / or medical student (HE – always supervised by LE). Interviews with inpatients took place on the ward where they were being treated, either at the bedside or in a separate room. An interview with a discharged patient was conducted over the telephone. Staff interviews occurred in a variety of settings and formats, including face-to-face in the workplace, remotely via Microsoft Teams or by telephone. All interviews were digitally recorded and transcribed verbatim.

### Qualitative data analysis

Qualitative data analysis used systematic coding and recoding as an iterative process by FM and LE. The data package, Nvivo v.12 helped to organise the data and identify key sub-themes.

### Quantitative Questionnaire data collection

A bespoke questionnaire was devised to gather information about individuals’ experiences, perceptions and opinions of sound and noise on the unit where they were currently working or being treated (available in supplementary information). It included multiple choice questions, Likert-style responses and free text responses. We were granted permission to use the validated, reliable(13) Weinstein Noise sensitivity scale(14) to assess noise sensitivity across participants. This is a 21-item scale with items scored from 0-5 (strongly agree through to strongly disagree, or vice versa with some negative scoring. Missing answers were imputed by calculating the mean score of answered questions and adding multiples of this according to the number of answers missing, up to a maximum of 10 missing questions.

### Quantitative questionnaire data analysis and findings

Questionnaires were analysed using Excel and GraphPad Prism. As this study was not statistically powered for specific findings, most of the analysis was simple and descriptive, with Kruskal-Wallis or Mann-Whitney tests used to compare between more than two or two groups respectively, and Fisher’s exact test to compare categorical data. Linear analysis for the Weinstein noise sensitivity scale was carried out in “R”.

### Merging of qualitative and quantitative data

The team met regularly to discuss emergent findings. A convergent parallel design (15) to the data collection and analysis (figure 1) was adopted to enable the mixed data to be considered with parity and to generate more encompassing understanding of hospital soundscapes.

### Reporting

Data are reported in accordance with Mixed Methods Reporting in Rehabilitation and Health Sciences(16).

## Results Sound levels

As shown in table 2, all sound measurement levels exceeded those recommended by the WHO, across all settings and time of day. Even in the quietest unit (NRU), both daytime and nighttime LA_eq_ would be perceived as being considerably louder than that required by the WHO guidelines (Day - 54.7 vs 35dB(A); Night - 43.6 vs 30dB(A)). These results suggests that there is little in the way of aural respite in most units.

**Table 2:**
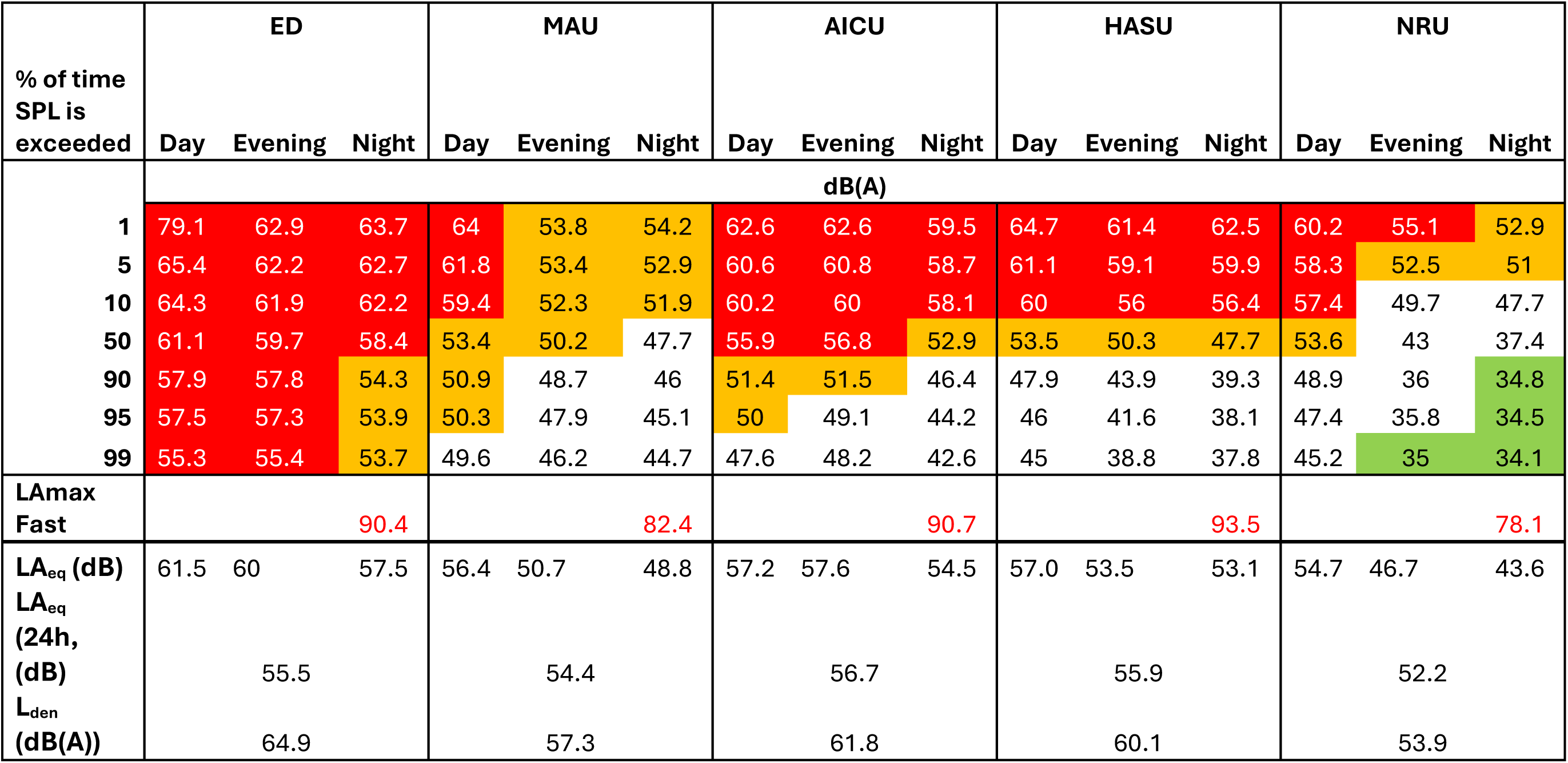
Upper section represents sound level measurements across units. Green indicates sound at a level conducive to sleep (</=35dBA); red indicates sound that could cause “serious annoyance” (>55dBA); orange “moderate annoyance” (50-55dBA). ED = Emergency Department; MAU = Medical Assessment Unit; AICU = Adult Intensive Care Unit; HASU = Hyperacute Stroke Unit; NRU = Neurorehabilitation Unit. Lower section represents global sound pressure levels recorded over 7-day periods in each unit. Note WHO recommends that LA_eq_ day (dB) and LA_eq_ night (dB) should not exceed 35dB(A) and 30dB(A) respectively.

### Annoyance and sleep

The WHO thresholds for moderate annoyance, that is where sound pressure levels exceed 50dB(A) would be experienced

- 99% of the night – ED
- 95% of the day – MAU and AICU
- 90% of the evening – AICU
- 50% of the day, evening and night – HASU, day – NRU, evening – MAU, night – AICU
- 10% of the night – MAU
- 5% of the evening and night – NRU

The WHO thresholds for serious annoyance, that is where these sound pressure levels exceeded 55dB(A) would be experienced

- 99% of the day and evening – ED
- 50% of the day and evening – AICU, night – ED
- 10% of the day, evening and night – HASU, day – MAU and NRU, night – AICU
- 1% of the evening – NRU

The WHO threshold levels compatible with achieving a “reasonable” sleep quality (<35dB(A) overnight, with an LA_Max_ for a single event <45dB(A)) would be unlikely to be achieved in any of the departments studied, apart from a few hours overnight on the NRU.

Overall, the data suggest that in the main, units were unlikely to be conducive towards rest or recuperation and may give rise to an increased potential for annoyance than would otherwise be experienced had the units complied with the recommendations made by the WHO.

### Participants

In total, 23 staff members and 31 patients (17 with ABI and 14 without) were recruited across the five hospital settings (details available in supplementary online information).

### Soundscape experience questionnaire

Responses to questionnaires between patients with and without acquired brain injury were broadly similar.

- No staff felt that they could describe their unit as quiet, but 33% of patients (6 patients, spread across ED, MAU, HASU and NRU) felt that their unit was quiet.
- 50% of staff and 23% of patients felt that sound affected their wellbeing
- 86% of staff and 25% of patients felt that noise made it more difficult for them to communicate with people
- 59% of staff and 13% of patients reported that sound in the department made them feel annoyed.
- 80% of patients described sleeping better at home than in hospital and 48% were woken up more than once at night due to noise. Despite this, over 60% of patients felt that they were getting enough sleep and rest.

Respondents were asked if they had any suggestions that might improve the noise environment on the unit. 197 suggestions were made by staff and patients, with some duplication.

The most common suggestions were around

- providing soft close bins (suggested by 12 staff and 10 patients)
- providing noise awareness training (suggested by 11 staff and 1 patient)
- providing televisions or radios for patients (3 staff and 2 patients)
- moving equipment more quietly (e.g. bins or trolleys) (6 staff and 10 patients)

### Noise sensitivity

Analysis using a linear model showed that there was no significant difference in noise sensitivity by status as staff or patient (p=0.35), or by age (p=0.12), but that scores were significantly lower in men than women (p=0.01).

### Qualitative analysis

Themes were identified as people sounds, and material / equipment sounds. Subthemes were types of sounds, time and place of sounds, individual perceptions of sounds, responses to sounds and suggestions for improvement. We defined people sounds as including talking, shouting, singing, laughing, coughing, and bodily noises such as laboured breathing, fitting, and fluid loss (including vomiting). These were all sounds produced by people only and hence not derived from another source, such as equipment. Although equipment and other items are often operated by people, we focused on the impact of people sounds within the complex soundscapes of hospital settings for ease of analysis. Material sounds referred to the sounds generated primarily by equipment, such as metal bins, alarms, pumps, dishwashers, sluice machines, doors, radios, telephones, and personal protective equipment. While many of these sounds are invoked by people, they have been described separately for ease of understanding.

Qualitative findings are summarised in table 3.

**Table 3:** Themes, subthemes, examples of codes and interview excerpts. Abbreviations: ABI =acquired brain injury; AICU = Adult Intensive Care Unit; CD = compact disc; ED = Emergency Department; FFP = filtering facepiece; HASU = hyperacute stroke unit; MAU = Medical Assessment / Admissions Unit; NBI = non-ABI; NRU = neurorehabilitation unit; PC = personal computer; PPE = personal protective equipment.

| Thematic Category | Codes | Described as | Perception | Interview Excerpts |
| --- | --- | --- | --- | --- |
| **Conversational Sounds** | **Patients** rely upon conversation to make sense of their surroundings and attribute value judgements according to volume, content, and context of conversation. | Talking<br>Unintelligible<br>Disruptive<br>Orientating in time and place | Gossip<br>Unprofessional<br>Muffled<br>Panicked<br>Compromised safety. | “I don’t mind with patients, it’s when it’s with each other for a non-medical reason, chatting about what they’re doing at the weekend, I don’t like that.” Patient, ABI, MAU<br>“...the unnecessary giggling and shouting out between each other [staff], ...Hooting and laughing and ... I mean nobody wants the workplace to be unpleasant but you’re in a hospital, come on.” Patient, ABI, MAU |
|  | **Staff** awareness of escalating sounds can be disruptive to their working actions and how they respond to patients. | Talking is associated with the working environment and so inevitable. | Calm talking levels indicator of smooth running of the environment, well-being of patients and staff. | “...you know that a resus is being managed well because it’s actually quite quiet and there isn’t the flapping and the noise.” Staff, ED<br>“...or sometimes it can be very noisy and trying to listen to a chest, I can’t even hear a chest, what is going on here?” Staff, ED<br>“...you start with a noisy environment, that then makes people more agitated, they then become more noisy and it’s a spiral. And then you add the sensory impairment over and above that and it can be very noisy indeed.” Staff, MAU |
| **Bodily Sounds** | **Patients** find bodily noises inevitable in the hospital settings, but these sounds add to anxiety, disruptive sleep, and possible timely recovery.<br>**Staff** find disruptive patients attract more time and attention leading to tensions in working environment and | Shouting out<br>Coughing<br>Laboured breathing<br>Vomiting<br>Diarrhoea<br>Shouting, screaming, pacing, physically and verbally | Sudden deterioration/dying.<br>Uncontrolled emotions and distress<br>Bodily sounds of concern identified as persistently disruptive, violent, | “...but it is distressing to hear, yeah. It makes you realise that you’re in a hospital when you can hear people being ill, do you know what I mean?” Patient, ABI, MAU<br>“...screaming and shouting, I will always go and investigate. Partly because I like to de-escalate. Well, I think it unsettles ... I mean proper shouting, kick-off shouting, and kick-off screaming, the one that everybody goes ooh.” Staff, ED<br>“...a disruptive patient clearly can just cut through everything and affect the whole emotional climate of the place.” Staff, MAU |
|  | having to maintain emotional safety of others. | abusive, unreasonable. | and emotionally draining. |  |
| **Happy Talk** | **Staff** find positivity in talking with others, especially the normalising of events and progression of patient outcomes.<br>**Patients** found happy sounds as positive and affirming, providing sense of being cared for and with others nearby. | Singing<br>Chatting<br>Jokes<br>Inclusion<br>Reassurance<br>Sharing positive news<br>Shift changes<br>Reassuring,<br>Pleasant, hopeful, human, being together. | Patient talking as validation of health improvement.<br>Genuine human connection.<br>Inclusive.<br>Orientation to time of day and regular events.<br>Health improvement.<br>Normalising the working day. | “I love hearing when a patient’s had their trachy [tracheostomy] out and I can hear their voice. There was one lady that I looked after and she had a trachy in for quite a while and I wasn’t looking after her this particular day but I walked past and she went ‘Hello’, I went ‘Oh ...’, aw that’s great, I love that (laughs).” Staff, AICU<br>“...the human interaction, the laughter you know, people being good-humoured, jokes...those noises are kind of genuine kind of human interaction and well they’re very humanising in a setting where it’s often possible to be very medical and non-human or inhuman about how you do your communications.” Staff, MAU<br>“I do like a little bit of noise. It’s like when I have the radio on at home you know, or the television, it’s a bit of background noise that I do like because I like to know there’s somebody else out there...” Patient, ABI, MAU<br>“... the doctor came to me and I’d had a scan and he says it hasn’t grown, your lump hasn’t<br>grown in your head, so that was a nice one (laughs).” Patient, ABI, MAU |
| **Night Time Sounds** | **Patients** find night time conversations as disruptive to their sleep, isolating and anxiety provoking. Night time talking and verbalisation can provide reassurance of staff being present.<br>**Staff** are aware of patient distress at night but they do not readily distinguish between day or night in their working activities | Hooting<br>Loudness<br>Inappropriate discussions<br>Working<br>Discussions<br>Gossip<br>Social catch ups with others<br>Chatting to patients in need | Disruption<br>Intermittent<br>Worrying<br>Sound Sensitivity<br>Isolation<br>Job necessity<br>Wellbeing and smooth working between staff<br>Ignorance of patient needs for sleep and recovery. | “Yeah, you don’t really want the hooting. The lady opposite me, I think she must have a bit of dementia and she just suddenly ... I was sleeping and she just suddenly shouted out a kind of urgh.” Patient, ABI, MAU<br>“Normally when I have a seizure I go to sleep for a while and then that helps me recover. Because I’ve got this really bad headache and it’s not helping because it’s too warm in here ... am pretty sensitive to noise anyway, that seems to be how my hearing works. Loud noises just goes straight through me.” Patient, ABI, MAU<br>“I’ve been really appalled actually at staff lack of care and insight into the fact that people are trying to sleep.” Staff, MAU<br>“As a patient I would say this is ... and you quite often hear people at night in hospital say I’ve had no sleep. And it’s not the patients, it’s the staff. Because it’s their job.” Staff, ED |
| **Equipment Sounds** | **Metal Bins** are a major source of | Clanging<br>Banging | Disruptive<br>Intrusive | “It’s a three-clang thing isn’t it because your foot goes on the thing ...it hits the wall and then |
|  | sporadic disruptive sounds which impact on patient and staff wellbeing. | Reverberation<br>Echo | Irritating<br>Anticipatory<br>Frustrating | it slams back down again. And it's a massive bang." Staff, MAU<br>"the bins are so loud (laughs) and it really makes ... it makes me jump sometimes but it also sometimes makes people, makes the patients jump." Staff, NRU |
|  | **Alarms** were in use in most contexts, multiple alarms in use in the acute areas such as intensive care.<br>**Patients** found them tricky to assimilate to their use and so produced mixed reactions from patients who concomitantly regarded them as reassuring, anxiety provoking and inevitably signifying a loss of control and acute illness. | Bleeps<br>Bongs<br>Tick Tock | Comforting<br>Persistent<br>Annoying<br>Irrelevant<br>Pointless<br>Loss of control<br>Uncontrolled<br>Weird | "I didn't get why it had to bleep, I really didn't. And I heard somebody else complaining, well not complaining but talking to relatives saying about the bloody bleeper thing. One's bleeping, then another one's bleeping. Don't know why they have to have a sound?" Patient, ABI, NRU |
|  | **Staff** generally found alarms as an inevitable part of their working environment. They questioned their effectiveness in timely responsiveness and frequently cited them as failing, disruptive and inconsequential. | Bleeping<br>Tick Tock<br>Continuous<br>Loud | Antiquated<br>Disruptive<br>Persistent<br>Stress inducing<br>Zoning out<br>Uncomfortable | “but it was beeping, then it was going like tick-tick-tick as well at you and it was like you’re not only alarming, you’re making these weird sounds. And it was like ... and I was going right, where’s the off button because I wanted to try and see if I could reboot it and couldn’t even find that. So I was getting angry because I couldn’t find that (laughs).” Staff, AICU<br>“you’ve pulled and pushed every single knob, button, tube you can and it’s still alarming. And you get someone else to tell you and they don’t know and it’s just going on and on and on and on. And you’re going for crying out loud. It was really literally doing my head in yesterday because none of us could figure out why it wanted to alarm at us. I had that yesterday and I was like oh my God.” Staff, MAU<br>“I think the other day at one point I went ‘Can you stop your monitors?!’, because they were just bonging. And I think some people get so used to them going, they are observing them but<br>because they're there at the end of the bed and they're doing something else, they don't automatically go and stop it. I mean obviously, they're there for a reason but there's always a nurse at the end of the bed anyway, so I think ... but obviously, you can't turn them down just in case you are watching two patients and you've just nipped off for whatever reason. But it is very, very loud." Staff, AICU |
| **Personal Protective Equipment (PPE)** | PPE was in use and included masks, gloves, plastic aprons, gowns, extraction hoods, isolation units around bed.<br>**Patients** described the use of PPE as a barrier to communication, difficult to engage with others. They also worried about the risks of infection and the general isolation | Muffling<br>Dulling<br>Echo<br>Rebound<br>Whirring<br>Impossible to hear | Disruptive<br>Challenging<br>Irritating<br>Compromised<br>Concerns about understanding, infection control, | "Because all they could hear is the whir of the air cycling. So communication was really difficult because when you're working in full PPE, your face is covered and the normal surgical masks you can hear quite well but you underestimate how much you read facial expressions or lip-reading. And I think the masks made people deaf, just because you couldn't rely on that additional input. And with the PPE masks, it was like that and it just dulls the sound massively doesn't it?" Staff, AICU<br>"the current lot of FFP3 masks don't fit me either but I've chosen to wear one that almost fits rather than wear the respirator. Because |
|  | which PPE reminded them of.<br>**Staff** were frustrated with the continuous use of PPE and found communication difficult. Staff found the PPE itself as noisy and generating irritating sounds | Disorientating<br>Muffling<br>Challenging | Uncomfortable<br>Sound dampening | whilst I hate noise, it's still important to be able to hear properly and that does certainly make your job impossible wearing a respirator. You can't listen to chests (laughs), you can't because where do you put your stethoscope?" Staff, ED<br>"they've got sliding doors which make a noise in front of every patient, they have their own sliding door. And they sit inside a polythene room. And so obviously there's less cross-pollination between patients because the polyethylene acts as a little bit of a sound barrier. But when you're in it with the patient it's very echoey inside the room. And I think again that might add to delirium and the sort of sense of being sensorily overwhelmed by your setting." Staff MAU |
| **Conflicting Sounds** | Clusters of sounds which identified as positively welcome sounds to participants and also negatively | Squeaky wheels (trolleys)<br>Clattering (refreshments)<br>Music (radio)<br>Chatter | Welcoming<br>Orientation to time<br>Normalising<br>Conflicting<br>Annoying<br>Subjective sounds | "Yes, the tea trolley makes me happy. We can have cake in kids, nobody complains. We have the tea trolley, nobody complains. We've never ever had a complaint from a family." Staff, ED |
|  | received by others in the same settings.<br>**Patients** valued routine sounds as providing a sense of normality and orientation in location, time and place.<br>**Staff** valued the “background” of normalising sounds in attempts to make the workplace more informal and relaxed to work in. However, some staff found music to be subjective and at times a distraction from maintaining focus. |  | Routine, Relief, Reassurance. | “You get used to the lady coming ... you get used to I call it the chuck wagon coming along, you know when your dinner’s coming or the tea or whatever. Don’t oil the wheel on the chuck wagon because I need to know that’s coming.” Patient, NBI HASU<br>“And sometimes like we try and have radios playing quite a bit. I think the radios are a bit of a tricky one though because some people can’t always say oh I don’t want to listen to that at the moment. And I think sometimes it’s assumed and perhaps they’re left playing with the same CD that they’ve had on for the past three days. And they might be thinking oh God, not Tom Jones again.” Staff, NRU.<br>“I mean the nurses often play music through the PC. I actually always switch it off because they’re listening to crappy radio stations and<br>you know, I'm trying to do my ward rounds, so I just turn it off." Staff, MAU<br>"...when it's kind of culturally inappropriate but meaning in terms of age and demographic music, so actually what you've got is a bit of Kanye playing on Radio One and you've got a room full of 70-year-olds. That's really distracting. And again this raises, notches up the tension a little bit and I can never quite work out what that really is doing for anyone?" Staff, MAU<br>"I quite like having the radio on" Staff, AICU |

## Discussion

This work has shown that hospital sound levels consistently surpassed WHO guidelines across all settings, but most markedly in ED. Interview and questionnaire responses indicate that sound and noise can have profound impacts on staff and patient wellbeing, mood and communication – both positive and negative. Patients with ABI exhibited both heightened and muted sensitivity to noise (as has previously been described in the literature(17, 18)), impacting their recovery experience. It is common for patients with ABI to have increased sleep requirements, including daytime sleep(19), indicating the need for a more nuanced understanding of hospital environments and sleep patterns.

Our work supports previous work showing consistently high sound levels across hospital departments(20). Prior studies have shown that lowering noise levels promotes reduced stress (physical and emotional) of staff and patients(21). Sound and noise can impair communication and understanding which is particularly important when sensitive clinical information may be being delivered (22), affecting identification and recall of key words(23), and, as staff participants in the study reported, their clinical interactions on ward rounds or inpatient consultations – “*the ward round is my operating theatre*” – where crucial decisions often need to be made quickly, and sensitive discussions conducted, which are both “*cognitively complex, it requires a lot of skill and brainwork*”. In areas such as ED, with fast-paced interventions and procedures, it becomes even more concerning – “*I think it’s particularly noisy, particularly for instance in a resus situation, noise is a real problem. And if you’re not heard effectively and you’re leading it could be dangerous actually.”,* confirming the associations between safety and effective communications.

“Voices” have been previously identified in other work as the most “bothersome” sounds in hospital, and the same study also found that patients tended to be less bothered by vocal sound than staff members(24) which is consistent with our findings.

Patients frequently were accepting that noise was necessary – “*It has to happen, so what can I do about it? They’re only doing their job, aren’t they*?” where staff identified that some, if not a majority, of staff noise, might be unnecessary in a clinical setting – “*staff are the noise-creators….go and talk somewhere else*!”

A range of interventions have been trialled with various levels of success in different institutions. Despite not having experience in acoustics, many of the participants in this study suggested interventions mirroring some of these, ranging from staff education, provision of sound level meters, enforcing “quiet times” on the ward, and estates and architectural modifications (24–31). A recurring theme in our work which did not seem to have been identified elsewhere, was around the noise created by waste bins opening and closing, which may have been exacerbated due to the increased use of personal protective equipment during the COVID-19 pandemic.

### Strengths and limitations

Broadly the mixed method approach, utilising a convergent parallel design, was feasible within the complex setting of the hospital wards during the pandemic, allowing the integration and critical analysis of data across several disciplines. We recruited diverse participants across five departments, allowing us to explore a range of potential contributors to hospital sounds and their perception. However, sample sizes were limited and there was likely self-selection bias among staff and prospective patient sampling may have resulted in the exclusion of potentially useful contributions by patients who may have identified differing perspectives.

Some differences in the expectation and experience of hospital sounds were identified between participants, with less tolerance among staff participants than patients. We had anticipated seeing more of an impact of hospital sounds on patients with ABI than patients without ABI. We know that many patients are reluctant to complain about their care or environment whilst in hospital which may be one of the reasons why patients, even those known to be sound sensitive, may have given more moderate responses than anticipated by the research team. By default, the interviews and questionnaires could only be completed by patients capable of completing these, meaning that those with more severe communication or cognitive difficulties were excluded.

This research was conducted in a single hospital trust, so generalisability may be limited, although it is in broad agreement with previous research which shows that WHO guidelines are rarely, if ever, achieved(20) in hospital settings.

## Conclusions and suggestions for future work

This work has demonstrated the complexities of sensory environments in hospital settings, and cautions against assuming that all sound is negative. The WHO sound levels appear to be almost impossible to achieve in hospital settings, and so relevant noise standards may need to be reassessed. We propose that attention needs to be given towards enhancing hospital soundscapes to improve patient sleep, experience and recovery and promote a healthy workplace for staff members which could improve staff retention in these pressured areas.

The role of PPE was more frequently mentioned in this work than in previous studies, likely due to its timing during the COVID-19 pandemic. As we continue to face risks of infectious diseases, the challenges to communication presented by PPE must be addressed.

Most participants offered at least one suggestion for improving the hospital sound environments. We plan to explore the ease, practicality and potential effectiveness of some of these interventions before developing implementation approaches with explicit measures of effectiveness.

## Data Availability

All data produced in the present study are available upon reasonable request to the authors

